# Six scenarios of non-medical interventions in the SARS-CoV-2 epidemic

**DOI:** 10.1101/2020.05.25.20112532

**Authors:** Peter Kempf

## Abstract

We investigate six scenarios spanning main parts of the decision space of non-medical interventions against the CoV-2 epidemic in Germany. Based on the notion of interventions-lifting we classify and evaluate the scenarios by five attributes (indicators): amount of interventions-lifting, death numbers, Public Health Care capacity, population immunity, peak dates of infections. For quantitative reasoning we use a simulated modified SEIR-model calibrated with actual data. We identify margins for intervention-liftings wrt. 13.05.2020 and discuss the relation to the effective reproduction number with a 6d-generation time. We show that, in order to constrain death numbers comparable to a strong Influenza epidemic, there is only a small corridor of 16% of possible liftings, with an additional 4% margin contributed by automated contact tracing. We show also that there is a much broader corridor of 50%+18%, though not overloading critical Public Health Care capacity, implying high death numbers.

## 1. Introduction

The worldwide outbreak of the covid-19 disease puts challenges on politicians and experts like epidemiologists, pharmacologists, virologists to design evaluate and implement adequate countermeasures to contain the pandemic. Reliable, *quantitative* epidemiological forecasts are necessary to validate far reaching decisions. To reduce the numerous degrees of freedom of the decision space, selected scenarios should be provided to decision makers.

Based on an enhanced simulated SEIR-model on a timely resolution of one day, we discuss five scenarios for Germany to react to the pandemic. We calibrate (*R*^2^ = 0.96) the model with actual data from 29. February to 13. May 2020 and available epidemiological data.

The next chapter introduces basic definitions of our SEIR model. Chapter 3 describes the calibration of the model with current data. Chapter 4 introduces four attributes (indicators) spanning a decision space of non-medical interventions.

We investigate the effect of *interventions-lifting*, i.e. the repeal of non-medical interventions against the spread of CoV-2, on all indicators. Furthermore the contribution of infection chain backtracking/contact tracing will be investigated. In Chapter 5 we consolidate the findings from Chapter 4 in six selected options: “Leave as is”, “Partial Lift-green”, “Partial Lift-yellow”,“Gradual Lift“, “Total Lift” and “Shutdown” to react to the pandemic. Chapter 6 introduces selected criteria, the scenarios can be evaluated against, and consolidates previous key findings into a decision matrix. Chapter 7 draws a short conclusion. The model equations are covered in the Appendix A. Some remarks to the used SEIR-simulation can be found in Appendix B.

## 2. The Model

We use an adopted version SEIR(**G**; *λ*, *τ*, *σ, ρ, ζ; κ*, *χ*, *η; α*, *θ*; *E*_0_*, I*_0_*, N*_0_*, R*_0_, var) of the well known SEIR-Model^1^ (see Fig. 1). The population is divided into several groups **G**.

**Fig. 1.**
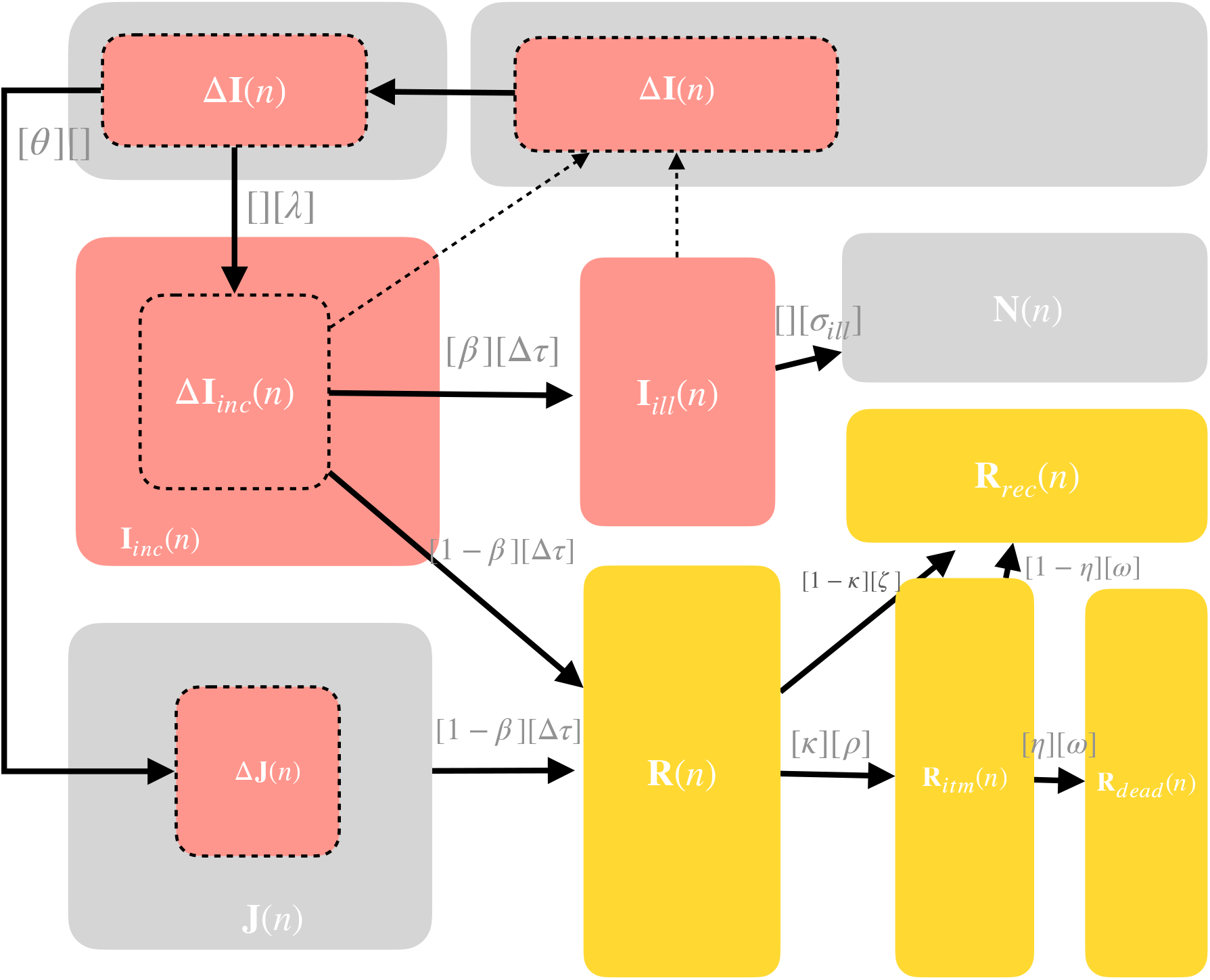
Modeling Scheme Colors indicate registered individuals (orange), infected or infectious individuals (red), transition coefficients are indicated as [probability][latency time].

Transitions between groups are indicated by probabilities and latency times.

Time is measured in days. Let *N*_0_ be the total population and *R*_0_ the *initial replication factor*. We define the *effective replication factor R_eff_ (n)* and the *effective replication rate r_eff_ (n) at time n* as

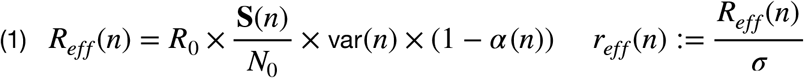

where **S**(*n*) is the number of susceptible individuals and var(*n*) models the seasonal variation, *σ* denotes the *infectiousity time and* the *intervention coefficients α(n):* 0..1 determine the strength of the interventions at time *n*.

Furthermore, we use the parameters from ^2^,^3^*: latency time λ, incubation time τ*, *time ρ from first symptoms to intense medical measures (ITM), ratio κ of ill individuals requiring ITM to registered cases*, the *death quote η* from ITM, the *dark figure χ* of undiscovered infections to registered cases with corresponding ratio 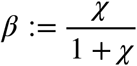, *recovery time ζ*, the *infectious incubation time Δτ:= τ − λ*. and the *infectiousity time σ_ill_:= σ* − Δ*τ from first symptoms*. To model the ratio of *identified* to all previously infected individuals by a registered individual (by contact tracing), we use coefficients *θ*(*n):* 0..1.

Each SEIR group **G** is modeled by a function **G**(*n*) mapping the current day *n* to the corresponding number of individuals defined as follows.

- *Susceptible* individuals **S**(*n*) which can be infected.
- *Exposed*, i.e. infected but not infectious individuals **E**(*n*).
- *Isolated*, therefore non infectious individuals **J**(*n*).
- Aggregated infected or *immune* (i.e. non susceptible) individuals **I**(*n*).
- Unregistered *infectious* individuals within incubation time **I***_inc_*(*n*).
- Unregistered *infectious* individuals with symptoms **I***_ill_*(*n*).
- Unregistered *recovered* individuals **N**(*n*).
- *Registered* individuals **R**(*n*).
- Registered individuals **R***_itm_*(*n)* requiring ITM.
- Registered recovered individuals **R***_rec_*(*n*).
- *Deaths* **R***_dead_*(*n)*.
- *Deaths* **D***_itu_*(*n)* caused by ITM bottlenecks.

Collected (measured) data between an time interval *n*.. *m* of group **G** is denoted as data[**G**](*n*.. *m*).

*Interventions-Lifting IL*(*α)* is defined as the *relative change in percent of the intervention strength α to the (calibrated) strength α*_0_*:= α* (*n*_0_) *=* 0.68 **in place at** *n_0_* = **13.05.2020**.

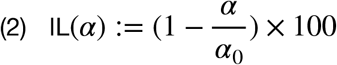

Hence, for a total lift of interventions IL(1) = 100% and for no lifting we have IL(0) = 0%. IL can be expressed in terms of *R_eff_* using (1) and **S**(*n*) ≈ **S**(*n*_0_) ≈ *N*_0_, var(*n*) ≈ var(*n*_0_)

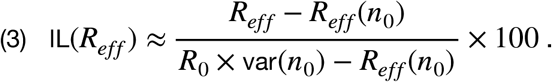

The *measured effective replication factor* 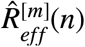 for generation time *m* at time *n* is derived from data of registered cases **R**(*n*) as^4^

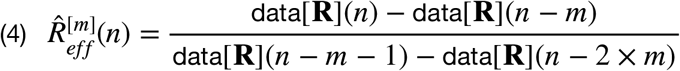

The *estimated effective replication factor* 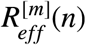 for generation time *m* at time *n* is derived from modeled registered cases **R**(*n*) as

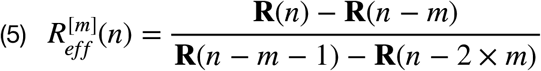

The *relative mean value deviation* 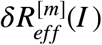 in a given interval *I* is defined as

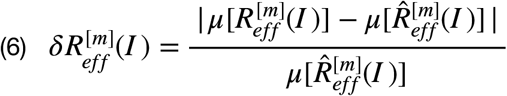

## 3. Simulation Calibration

Simulation range is nearly two years from 01.02.2020 to 31.12.2021.

We assume, similar to ^2^ and ^3^, the following values for the model parameters:

*N*_0_ = 8.0 × 107, *R*_0_ = 3.275; *λ* = 1; *τ* = 6, *σ* = 10; *ρ* = 6, *ζ* = 12; *κ* = 0.1; *ω* = 10; *η* = 0.5; *χ* = 4.0 with a 33%-seasonal dependency 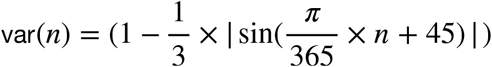. Furthermore we assume a basic immunity of 20% of the total population.

We have chosen a shorter latency time *λ* than in ^2^ and ^3^ as it explains better the rapid increase of infections at the beginning of the epidemic. Incubation time *τ* is a little bit higher (+0.5 d) but within limits, as the discrete simulation with a resolution of one day allows only integers in the difference equations. According to current data (from 13.05.20) the lethality ratio is 4.51 % of registered cases. This implies an ITM-ratio *κ* ten times higher than in^2^. We estimate (impute) initial values *I_inc_*:= **I***_inc_*(0) = 1000, *E*:= **E**(0) = 167, a dark figure ratio of 4:1 and calibrate the intervention coefficient *α* for three selected interventions at *n_schools_* = 16.03.20, *n_lockdown_*= 24.03.20 *n_lift_* = *n*_0_ = 27.04.20 to be *α*(*n_schools_*) = 0.1, *α*(*n_lockdown_)* = 0.68, *α*(*n_lift_*) = 0.68 = *α*_0_, so that the number of accumulated registered cases **R**(*n*) fits best (*R*^2^ = 0.963) to real accumulated registered cases data[**R**](*n*) from **01. February 2020 to 13. May 2020** (see Fig. 2) as collected by the CSSE^5^ from the Johns Hopkins University. The convergence of the estimated (red) and measured effective R-values with 6d-generation time is shown in Fig. 3. Within *I* = [05.04.20..13.05.20], we have a relative mean value deviation 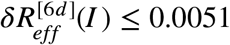, so we can assume 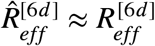. Interventions-lifting (3) reads with *R_eff_* (*n*_0_) = 0.57 and var(*n*_0_) = 0.9002:

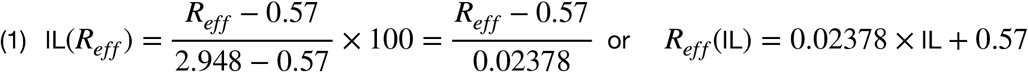

## 4. Indicators and Interventions-Lifting

For the description and discussion of the scenarios in Chapter 7 and 8 we will consider the following *indicators: peak date* of the resulting epidemic wave, the maximal number of *needed ITM/RU units*, the resulting degree of *population immunity* and the number of caused *deaths*.

Based on a set of simulations of the calibrated model defined in Chapter 2 and 3 for different ILs, we will investigate all indicators as a function of IL and will use the findings in the description of the scenarios in Chapter 5. Moreover we will evaluate the risk of an unintended secondary wave overloading Public Health Care capacity.

**Fig 2.**
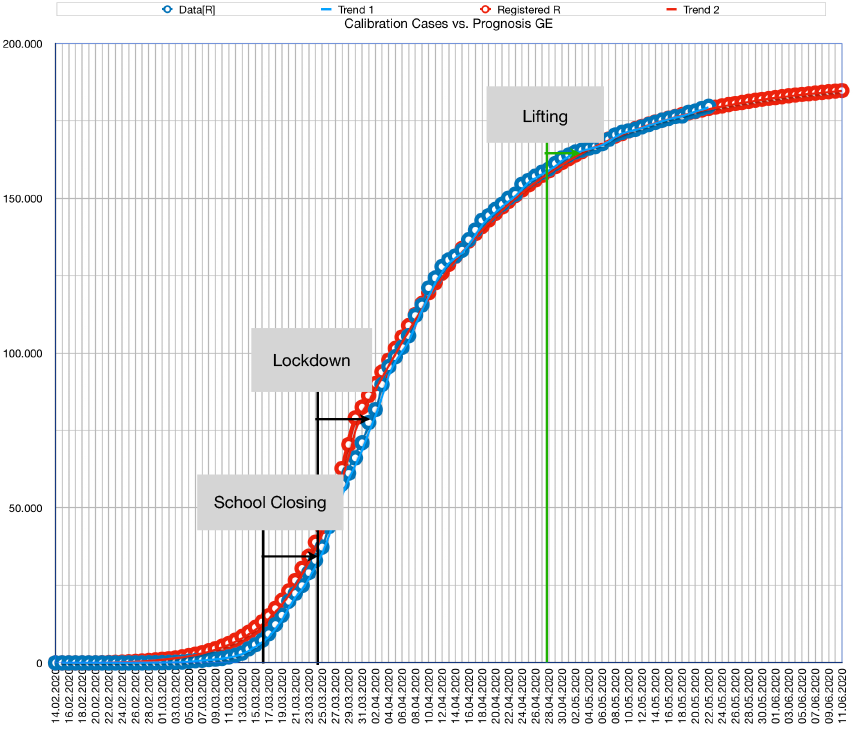
Calibration of forecast (red) with real data (blue) (R^2^ = 0.96)

**Fig 3.**
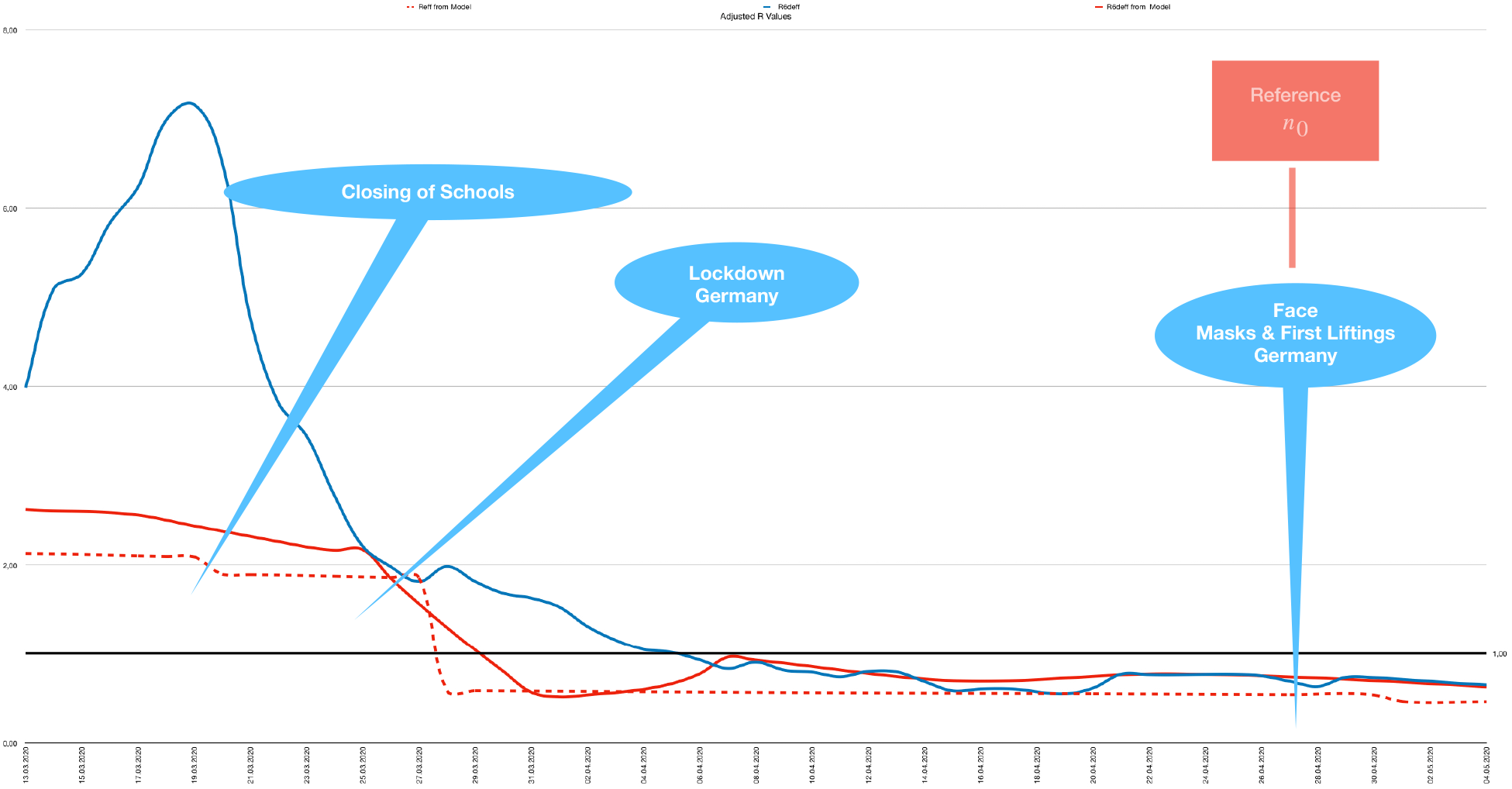
Modeled *R_eff_ (dotted red), estimated (red) and observed (blue)* 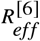 *R-values (6d-generation time)*.

**Fig. 4.**
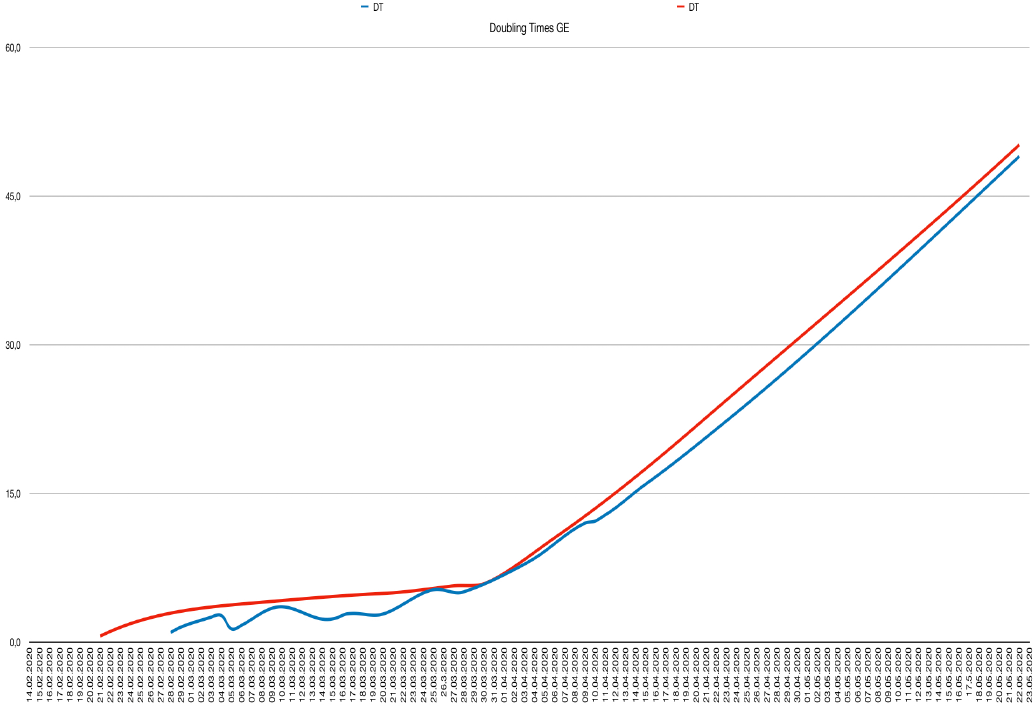
Doubling Times data (blue) and forecast (red)

### The Effects of Interventions-Lifting

Fig. 5 shows the required ITM units dependent on IL. Interestingly, the “ITM function” has a local minimum (“anomaly”) at 45%. There is a “yellow” corridor from **0% to 50%**, which is below free Public Health System capacity at 14.000 ITM/Respiratory Units^6^.

**Fig. 5.**
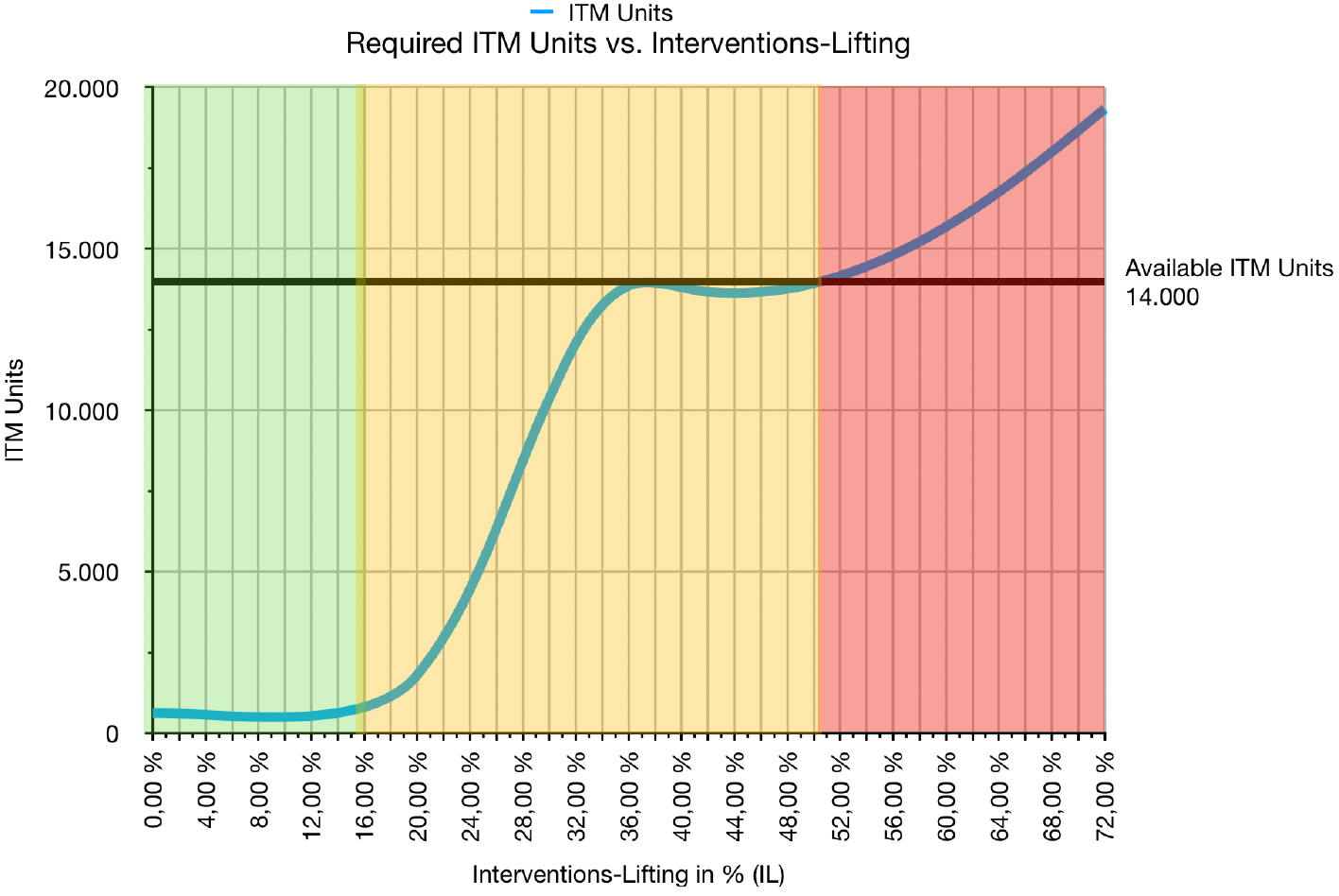
Required ITM units vs. lift of interventions. There is a corridor from 0%-16% (green) and from 0%-50% (yellow).

Fig. 9 shows a more narrow “green” corridor from **0% to 16%** where the number of deaths is in the range expected of a strong Influenza epidemic.

Fig. 6 shows the ITM and death *rates* wrt. percentage of interventions-lifting with higher values at the 16%-edge of the “green” corridor. At this edge there is some risk of ending up with higher death numbers than expected.

**Fig. 6.**
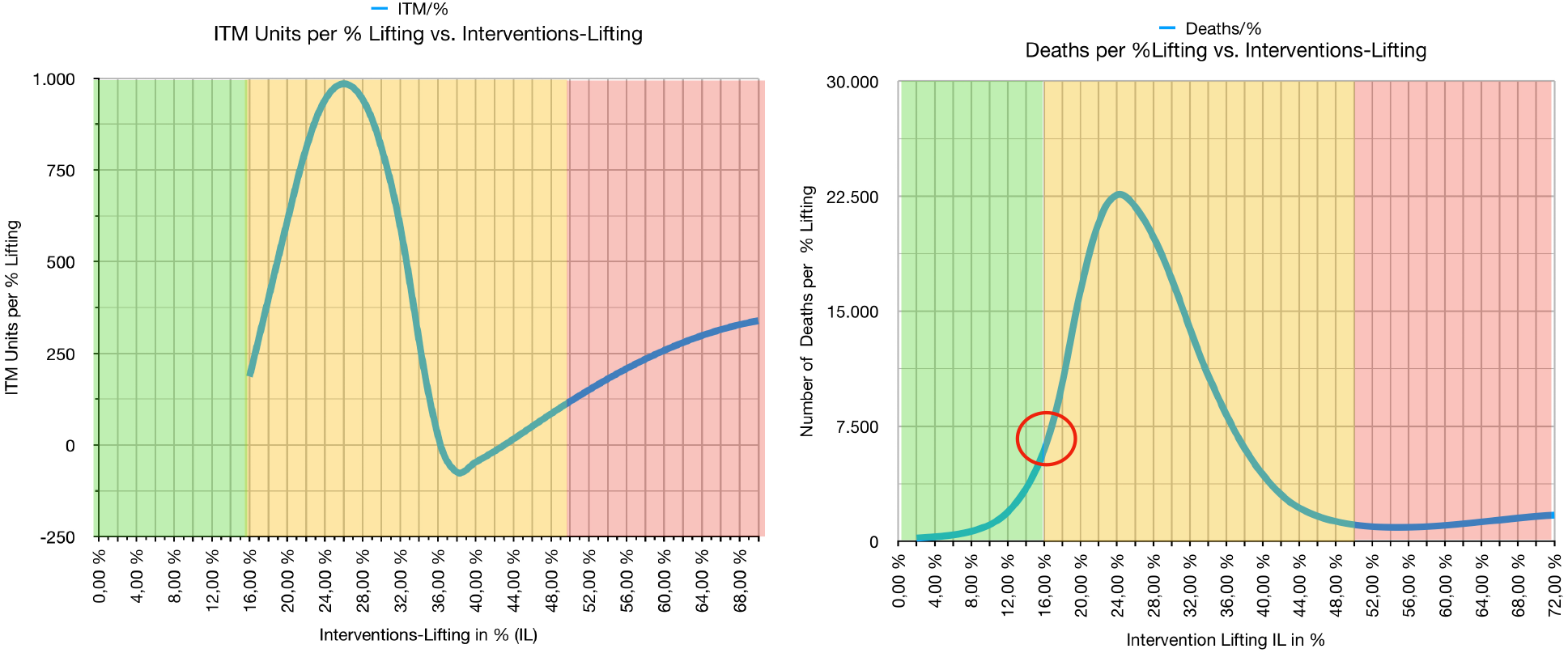
Sensitivity of interventions-lifting. Left diagram: margin of needed ITM units per % lifting. Right diagram: margin of deaths per % lifting.. The red circle indicates the higher death rate at the border of the green and yellow corridors.

Fig. 7 shows the dates where the epidemic wave is peaking (maximum **I***_ill_*). For a “yellow” lifting of 50%, the peak will be expected in November 2020.

**Fig. 7.**
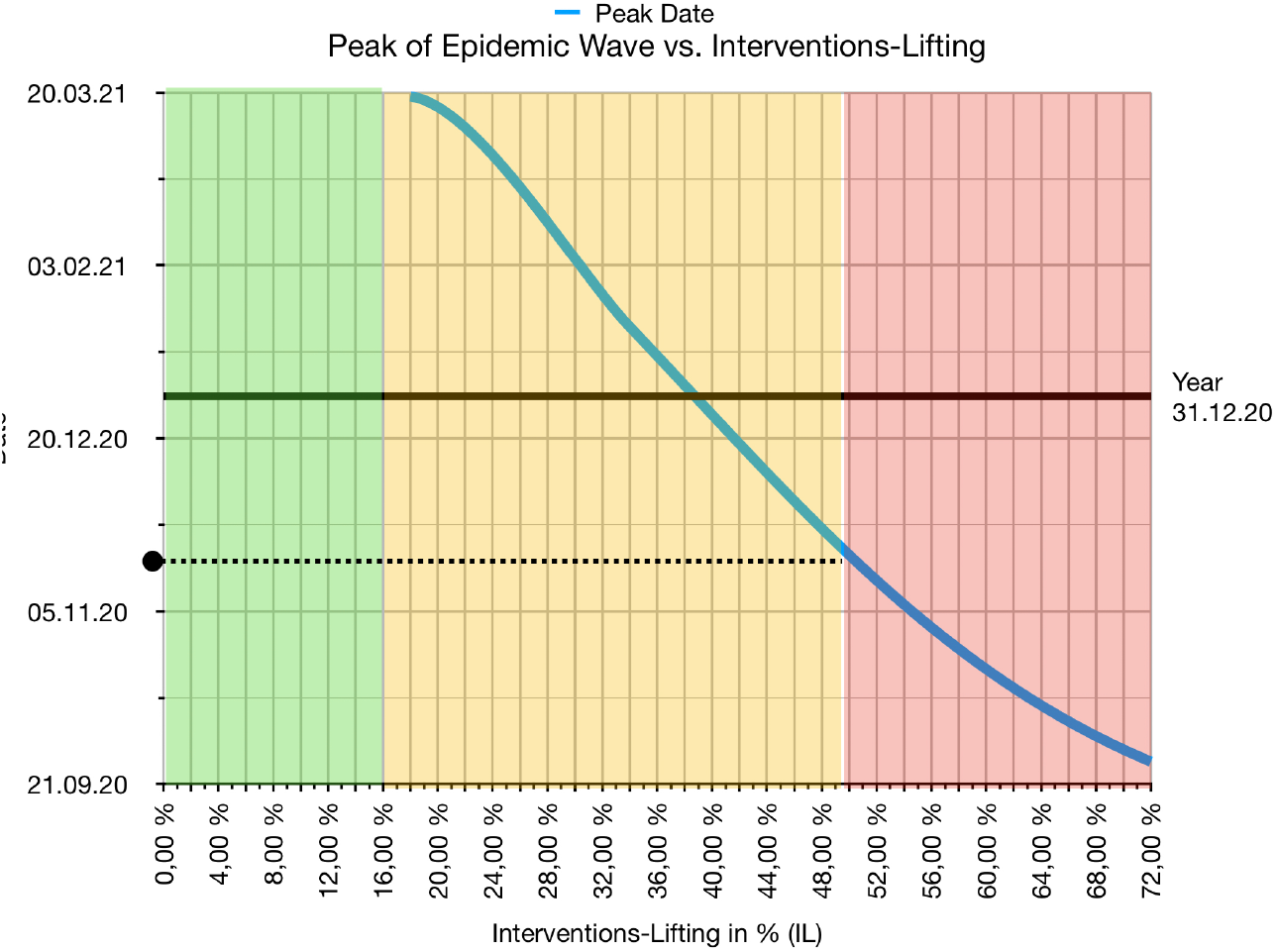
Peak dates of epidemic waves caused by interventions-lifting.

Dependent on the amount of interventions-lifting herd immunity might result. Fig. 8 shows the population immunity dependent on IL. At the right border of the “yellow” corridor herd immunity is achieved.

**Fig. 8.**
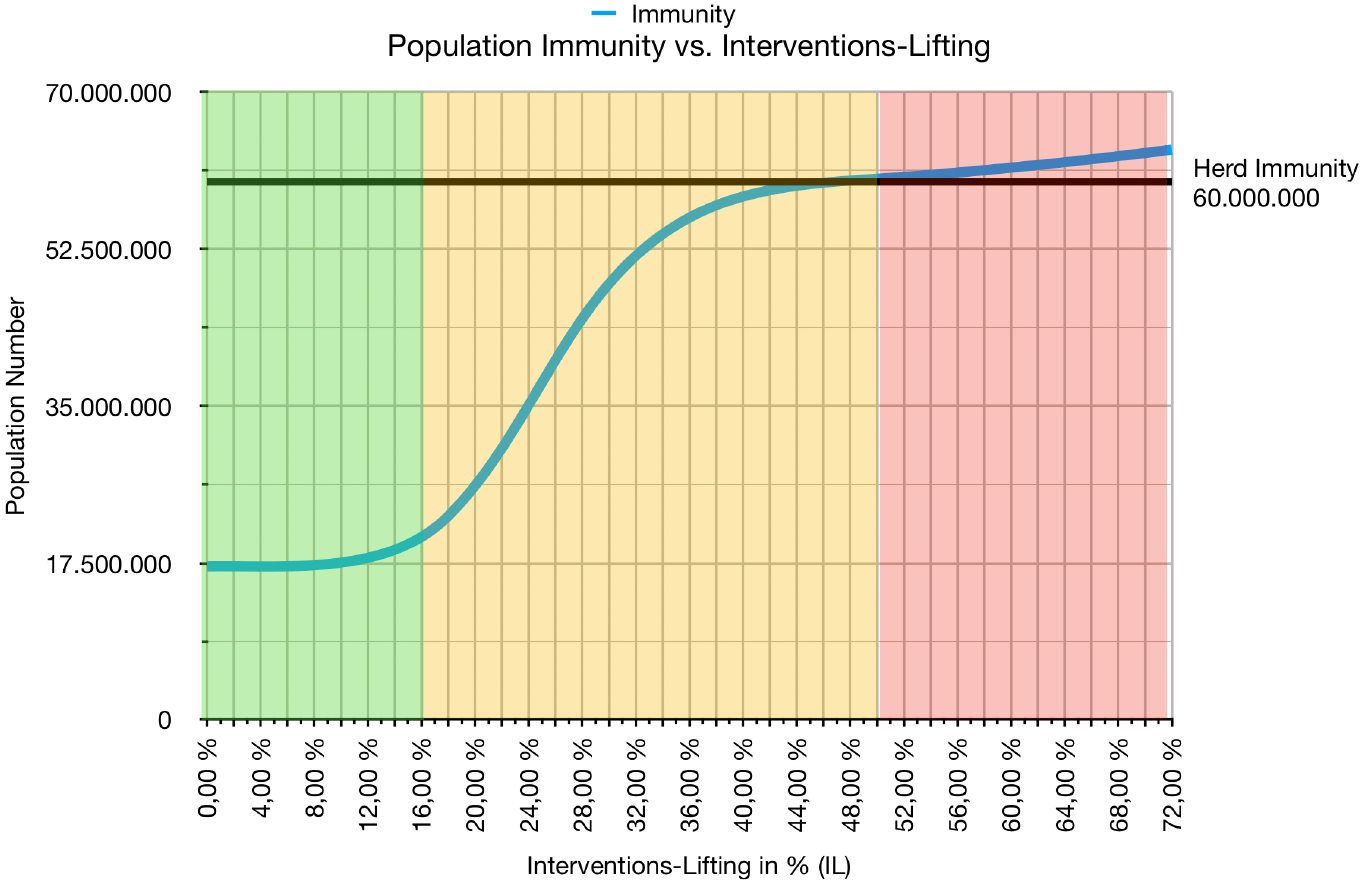
Population Immunity vs. Interventions/Lifting. At the right border of the yellow corridor herd immunity will be achieved.

The price for interventions-liftings achieving herd immunity is high. Even in the “yellow” corridor of Fig. 9 where ITM units may be sufficient, the number of deaths may be up to 380.000. Only the “green” corridor restricts the number of deaths comparable to a strong Influenza epidemic.

**Fig. 9.**
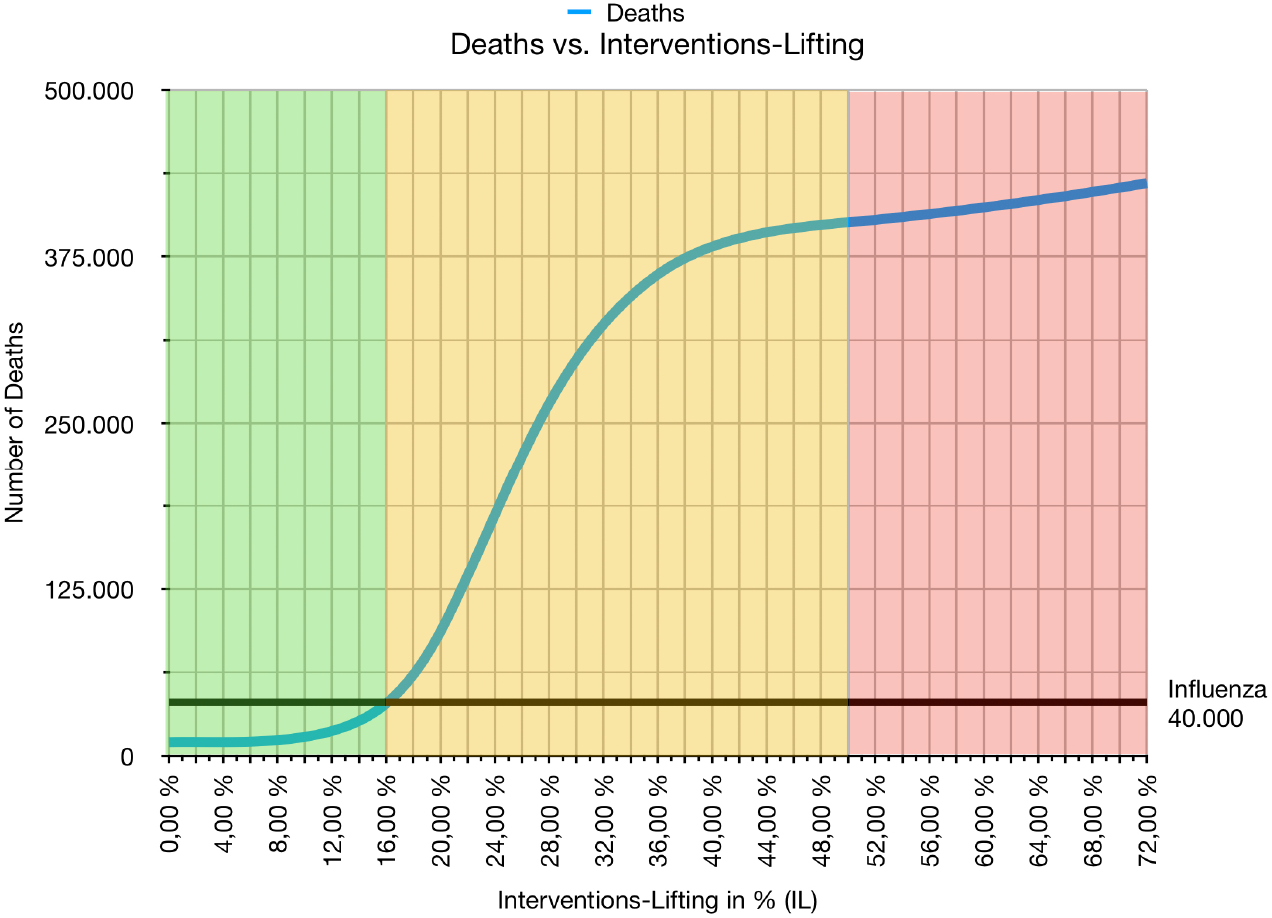
Deaths vs. Interventions-Lifting. High death numbers in the 33% corridor. Only the 20 %-corridor (green) has losses of life comparable to a strong Influenza.

For controlling the effects of interventions-lifting the knowledge of the *estimated and measured* effective *R*-value is essential. Fig. 10 shows the relation between IL and the *estimated* effective 6d *R*-value at start (*init)* and at the peak/end (*max, min)* of the epidemic wave.

**Fig. 10.**
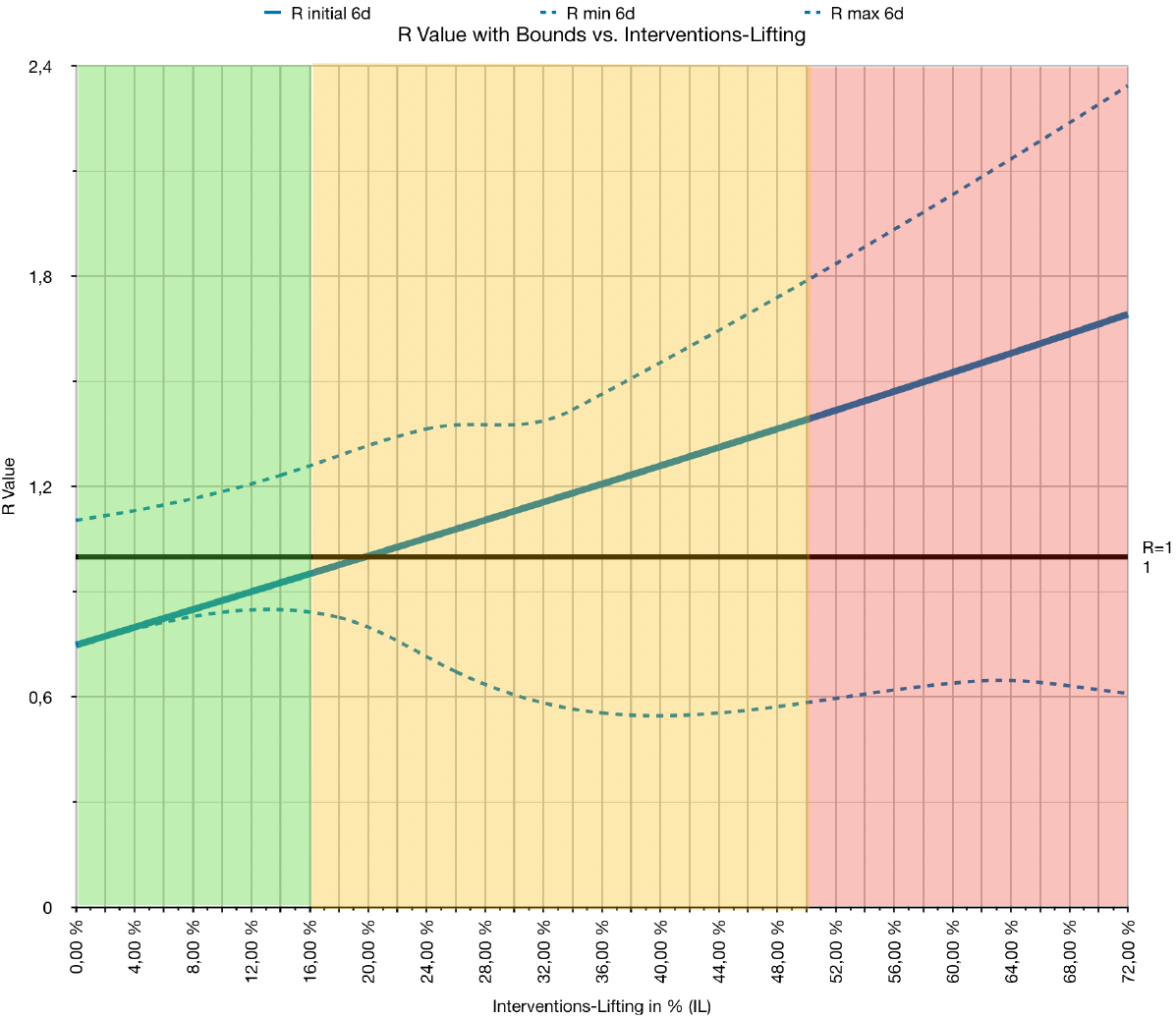
Calculated initial effective R-value (6d-generation time) dependent on interventions-lifting. R-value bounds of the resulting wave are indicated by dotted lines.

The simulations show a linear relation 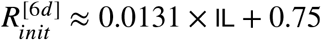. (Compared to (2) we have a *difference* 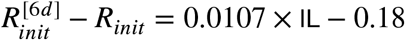 to the “real” effective R-value from 2.(1)). So, each percent of intervention-lifting changes the initial *R* by 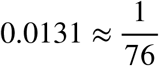. We thus estimate for a “green” 16%-lifting

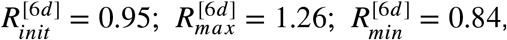

and for a “yellow” 50%-lifting

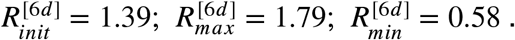

### Gradual Interventions-Lifting

Another promising option is the gradual lifting of interventions over a certain period of time just to stay below the trigger of an uncontrollable wave.

In the following we will investigate each indicator dependent of the percentage of gradual interventions-lifting on a **14 day period** starting from **beginning of June 2020 until beginning of September 2020** (seven intervention-liftings).

Fig.11 shows a small “green” corridor from **0 to 2.05%** (i.e. 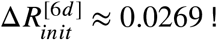) per 14d-interventions-lifting which has low death numbers and sufficient Public Health Care capacities. A wider extension (“yellow” corridor) to 4.34% (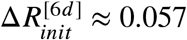) increases dead numbers dramatically, though not overloading Public Health Care. From 4.34% upwards, (“red” corridor), an uncontrollable wave will result.

**Fig. 11.**
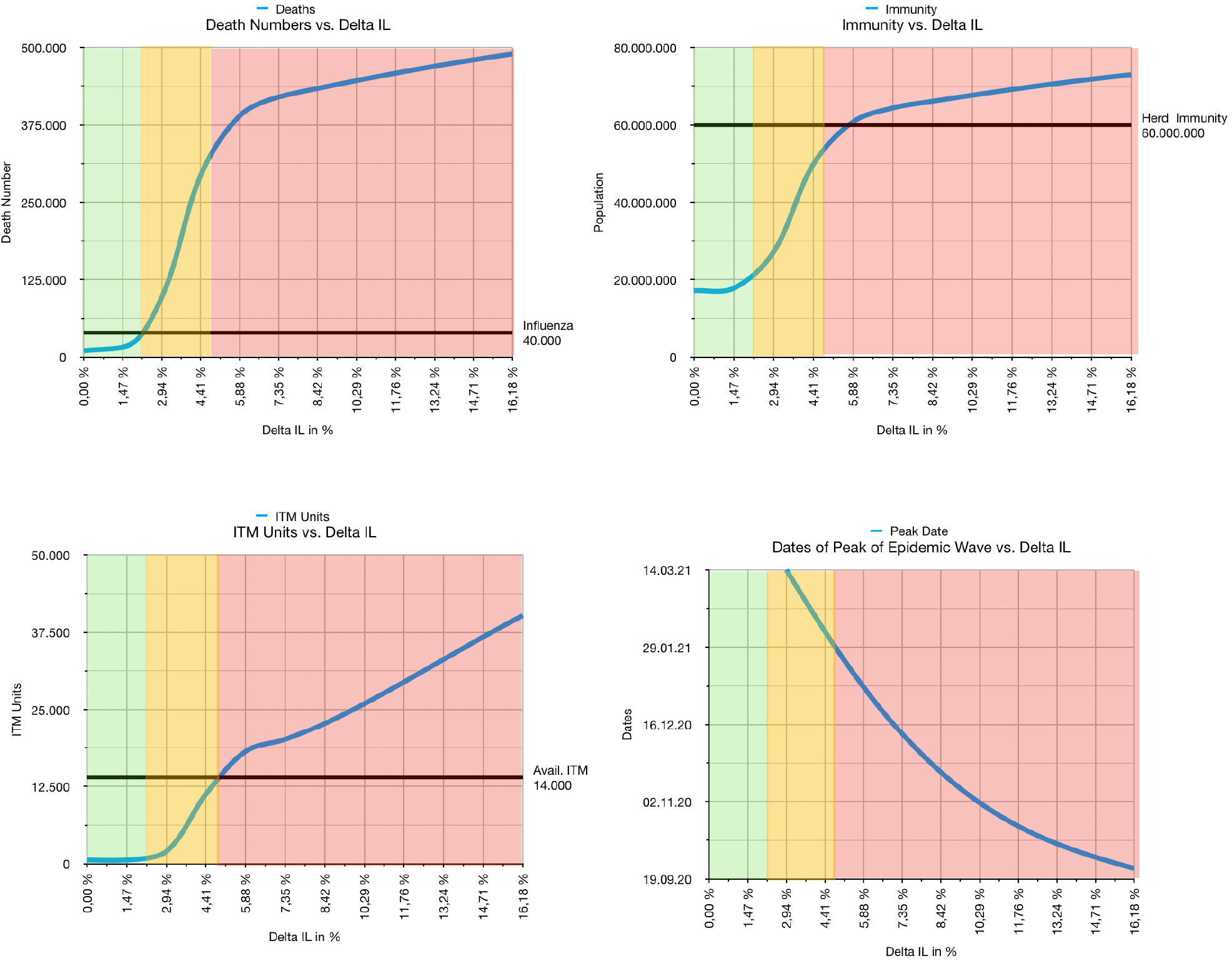
Indicators vs. 14d-gradual interventions -lifting. Colored areas indicate: low death numbers (green), high death number but sufficient ITMs (yellow), high death number and overload of Public Health Care (red).

The design of *a-priori* interventions-lifting measures which will meet the small corridor of 2.05% or 4.34 % may be challenging. On the other hand, with close control of the measured effective R-value, an adequate correction in the next gradual intervention may minimize the risk of an *R*-overshooting.

### Contact Tracing

What will be the effect of *contact tracing and isolating* individuals who have been potentially infected by a registered CoViD-19 positive tested individual ? We simulated the effect for a “green” 16%-lifting, assuming 100% of the immediately contacted individuals (one generation backtracking, say with mobile app) to be discovered.

Fig. 12 shows the effect of contact tracing on needed ITM units and number of deaths. The reduced numbers imply an additional margin of 3.3% of interventions-lifting in the “green” and, due to the anomaly of the ITM-function, 18% in the “yellow” corridor (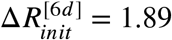).

**Fig. 12.**
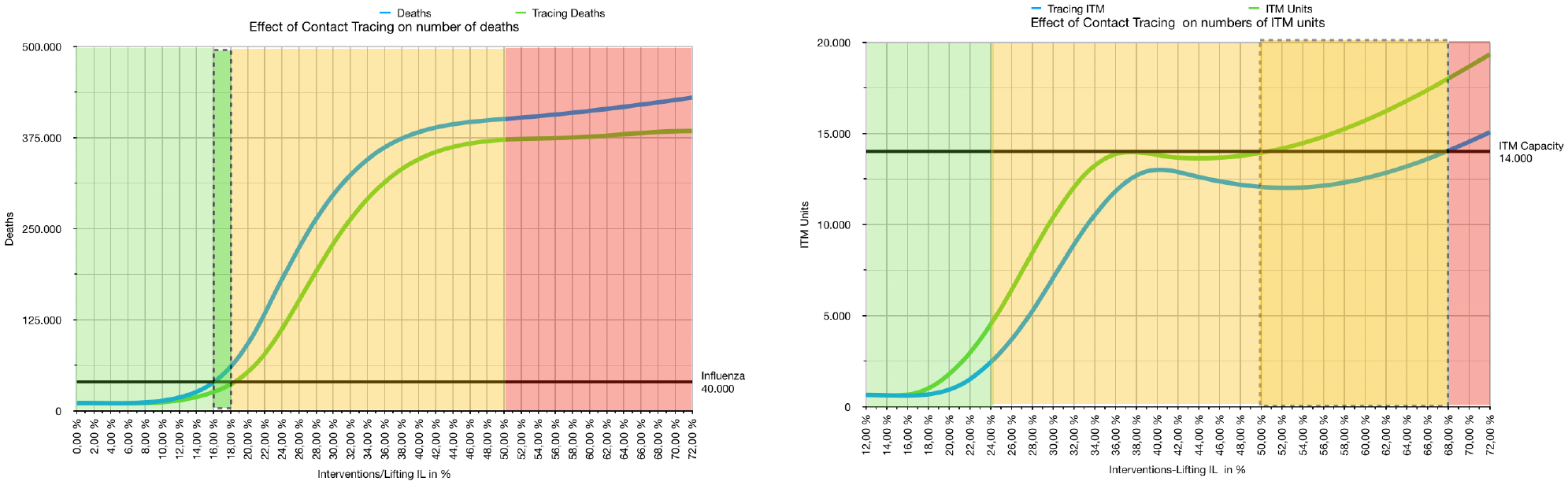
Effect of contact tracing on ITM units and number of deaths. Dashed areas show the additional margin of IL.

Even if the number of infected individuals is low, one-generation contact tracing will not prevent the emergence of an heavy epidemic wave resulting from a *total* interventions-lifting. Fig. 13 shows the severity of infection dynamics if interventions are completely repealed mid September 2020. Though the initial number of infections is low and aggregated isolations will be around 9.000.000 individuals, there is still a sharp and high peak of infections in mid November 2020. Main reasons are: the (assumed) insufficient contact tracing of only one generation, the (assumed) dark figure 4:1 of individuals which will be not registered and thus will not be traced, and the relatively long unidentified infectiousity with no symptoms during incubation time. Only if there is a *very* low number of infectious individuals, contact tracing may contain the infection dynamics.

**Fig. 13.**
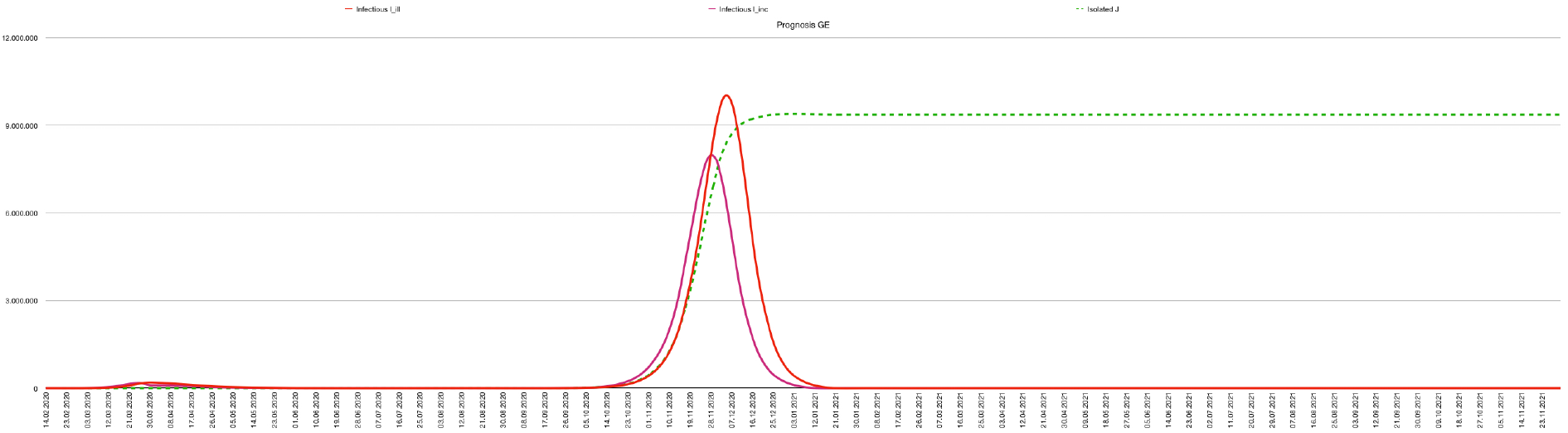
Effect of contact tracing on total Interventions Lifting due to incomplete extinction of the epidemic.

## 5. Scenarios

In the following we provide *five* scenarios covering relevant parts of the decision space of intervention-liftings that can be controlled politically. We assume a free capacity of 14.000 ITM units (incl. respiration equipment)^6^

### Scenario A “Leave As-Is”

Scenario A assumes the current intervention measures to be continued at the same level as of **13.05.2020** until end of 2021. Fig. 14 shows the resulting infection dynamics: The peak of infections has already been surpassed. Settle point of accumulated registered cases **R** will be beginning of July 2020 around 200.000. Expected deaths are 10.400. Herd immunity will not be reached. The peak load of the Public Health System has already been surpassed beginning April 2020 at 630 ITM units. Respiratory Equipment will be, by far, not exhausted.

**Fig. 14.**
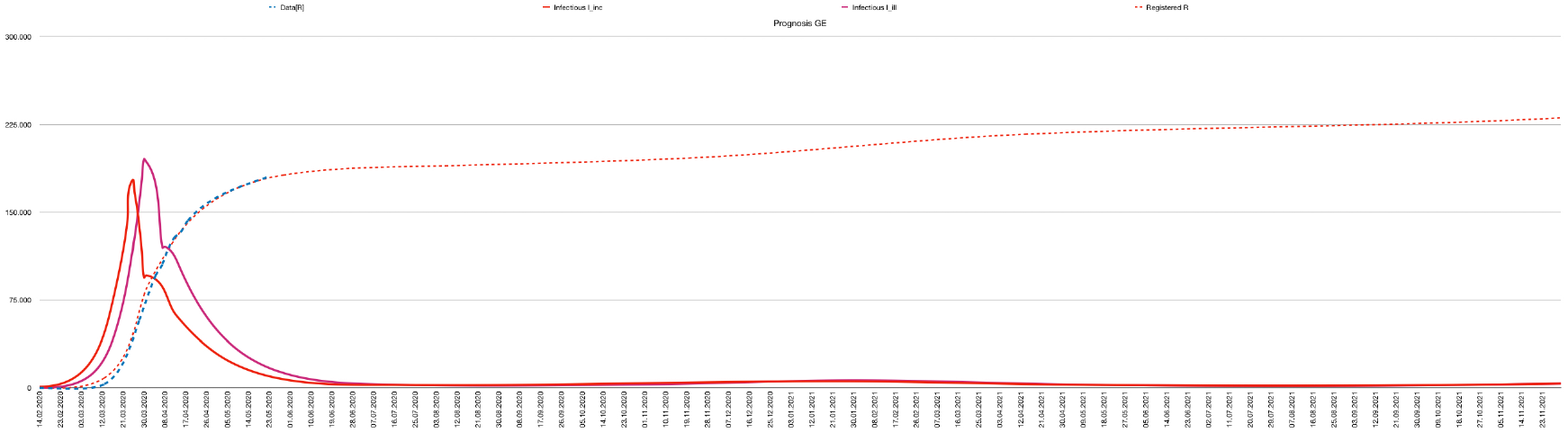
Infection dynamics of Scenario A. Number of infectious individuals during (red) and after (purple) incubation time. Registrations are indicated in dashed red, actual data in blue.

### Scenario B “Partial Lifting”

Scenario B assumes the interventions as of **13.05.2020** will stay till **end of May 2020**, followed by a maximal “green” repeal of 18 % (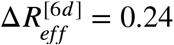), or maximal “yellow” repeal by 68 %, (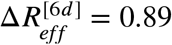) resulting in an epidemic wave peaking in March 2021 (October 2020). We assume contact tracing to be operational.

In the “green” wave 630 ITM units and in the “yellow” wave, 14.000 ITM units will be needed and thus, will not push the health system over limits (Fig.15). The “green” wave comes with a death quote similar to a strong Influenza epidemic (40.000), whereas the “yellow” wave will lead to 380.000 deaths ! Population immunity will be at 25 % at the end of the “green” wave and at 75 % at the end of the “yellow” wave.

**Fig. 15.**
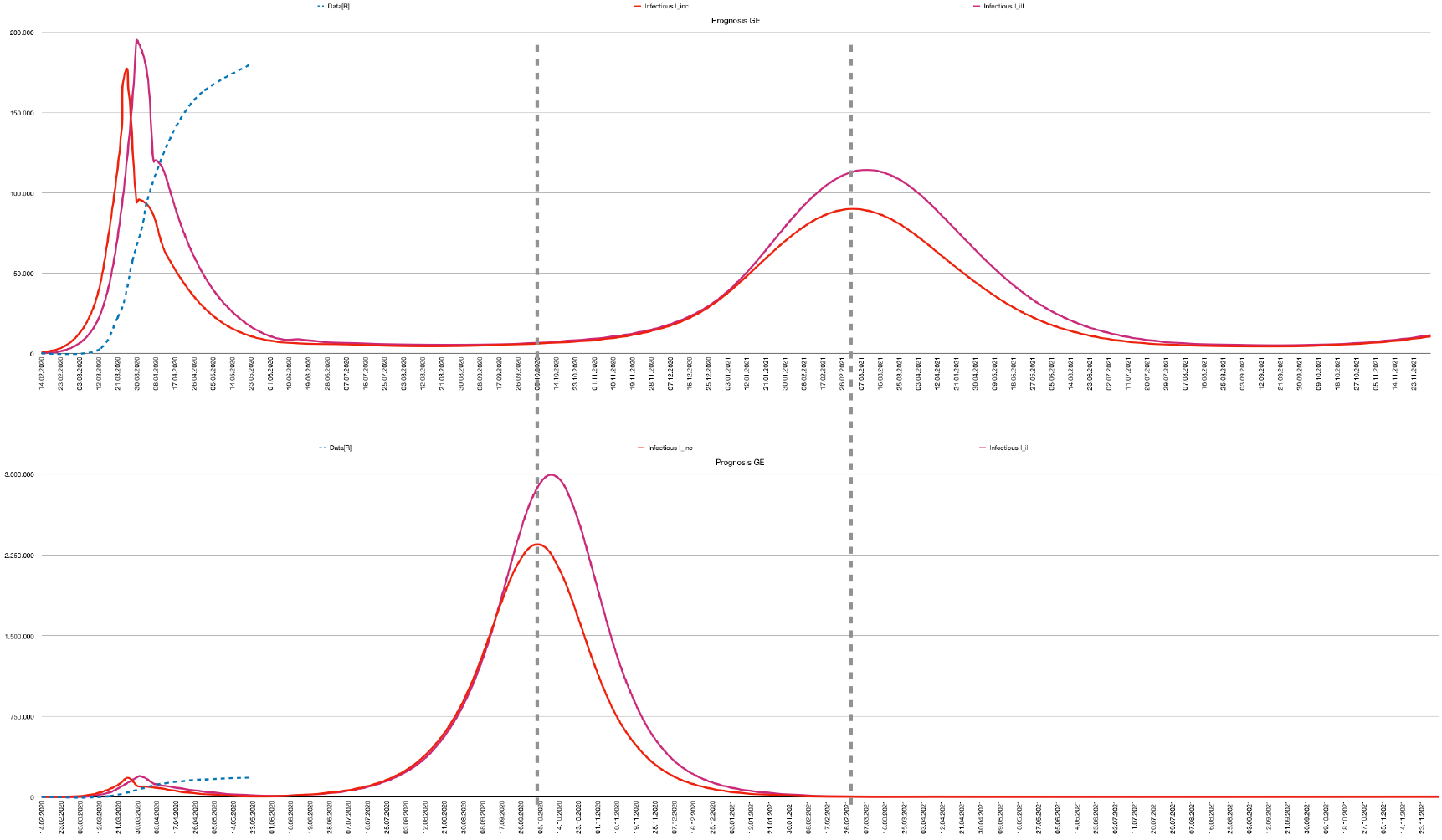
Infection dynamics in Scenario B. Waves of a 18% lifting (above) and of a 68% lifting (below).

### Scenario C “Gradual Lifting”

For Scenario C we assume maximal gradual IL-liftings in the “yellow” corridor of Fig. 11. Thus, ITM units will be sufficient, but the death number will be high. With operational contact tracing we have ILs by 4.34% + 0.47 % = 4.81 % (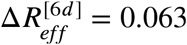) every 14 days starting from beginning of June 2020 until beginning of September 2020. This gives a total of 30.38% + 3.30 % = 33.68 %, (34.32 % less than in Scenario B !)

Fig.16 shows the resulting development of infections peaking in February 2021. The maximal required ITM units will be nearly 14.000. Public immunity will be at 67%, near herd immunity. Number of deaths will be at 330.000.

Interestingly, the corridors of partial and gradual lifting *do not* match. If interventions at the border of the *yellow* corridor of partial liftings at 50 % are split up in seven gradual liftings of 7,14% we end up in the *red* corridor of gradual liftings with a need of considerably more ITM units !

**Fig. 16.**
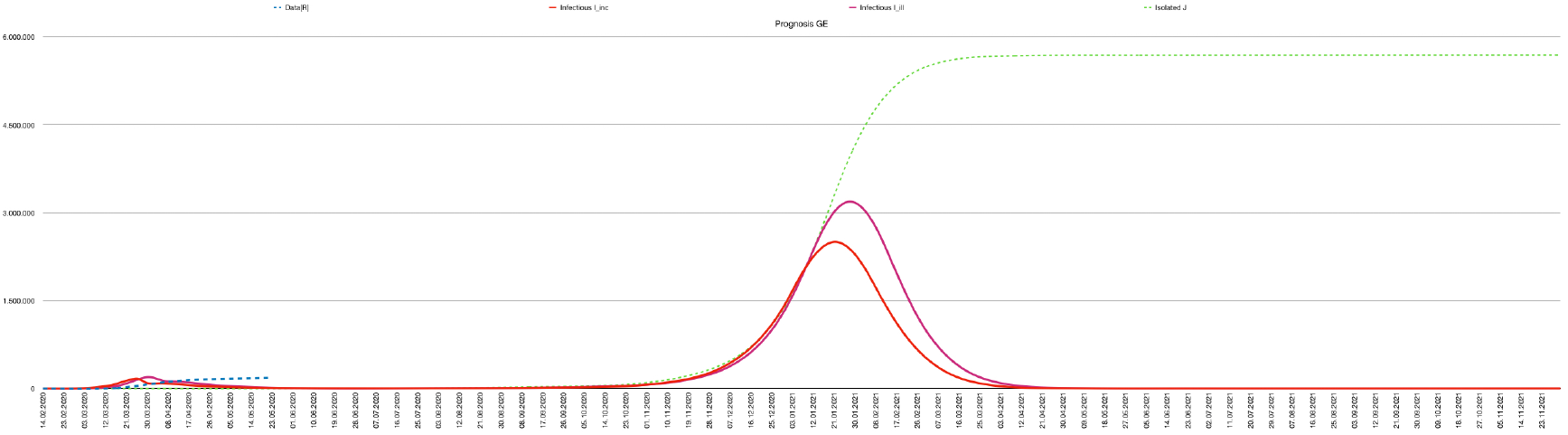
Epidemic wave in Scenario C with peak in January 2021. The dotted green curve shows the aggregated isolations due to contact tracing.

### Scenario D “Total Lifting”

Scenario D assumes the current interventions to be fully repealed in June 2020, followed by a massive epidemic wave in August 2020. The number of deaths will be about 630.000. Herd immunity will be reached immediately, at 86 %. The health system will be extremely overloaded, causing about 16.000 deaths due to ITM bottlenecks.

**Fig. 17.**
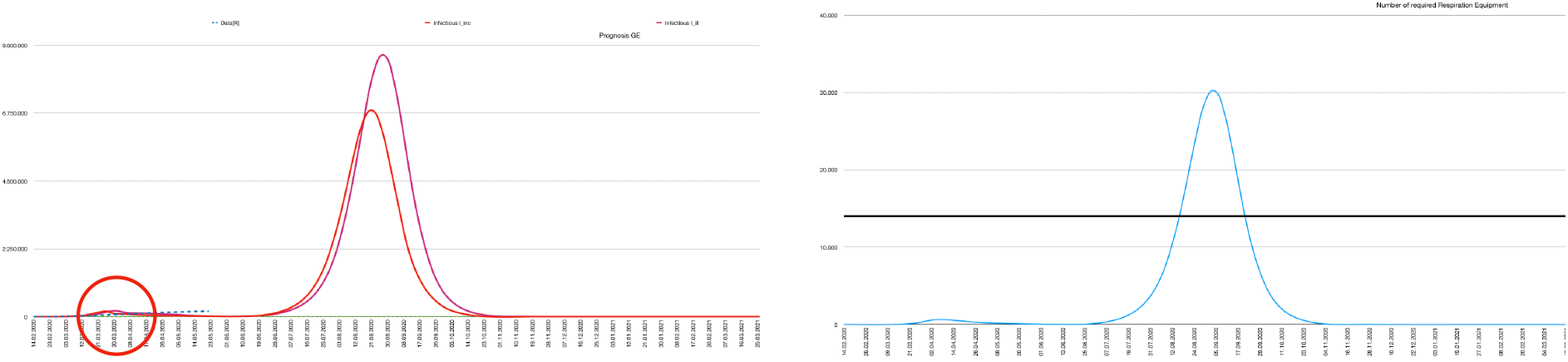
Scenario D: *massive epidemic wave peaking in August 2020. Infections (left), ITM units (right). For comparison: the current wave is shown within the red circle*.

### Scenario E “Shutdown”

Extreme social distancing measures for the whole population like total exit locks for **three weeks** (negative IL) will completely extinct the pandemic (Fig. 18). Workers in critical infrastructure will have to stay/live in their working area. Supply of the population will have to be guaranteed by government (armed forces, etc.). Total deaths will be below 10.000. Nevertheless, extinction is fragile as herd immunity will not be achieved and imported infections may cause a massive epidemic wave (Fig. 19).

**Fig. 18.**
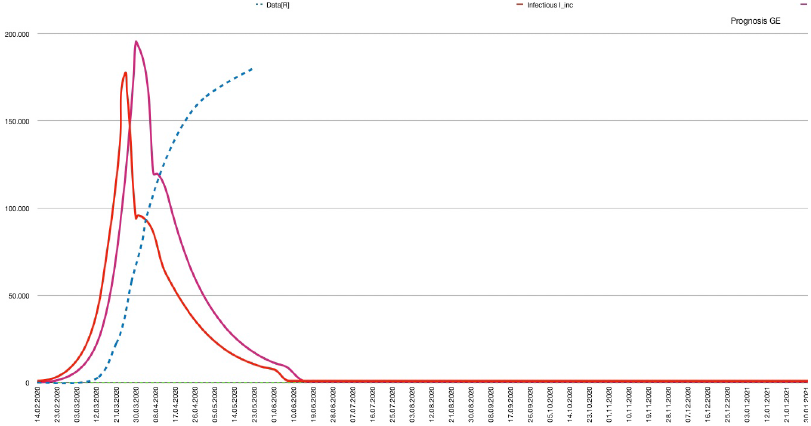
Extinction of Cases in Scenario E.

**Fig. 19.**
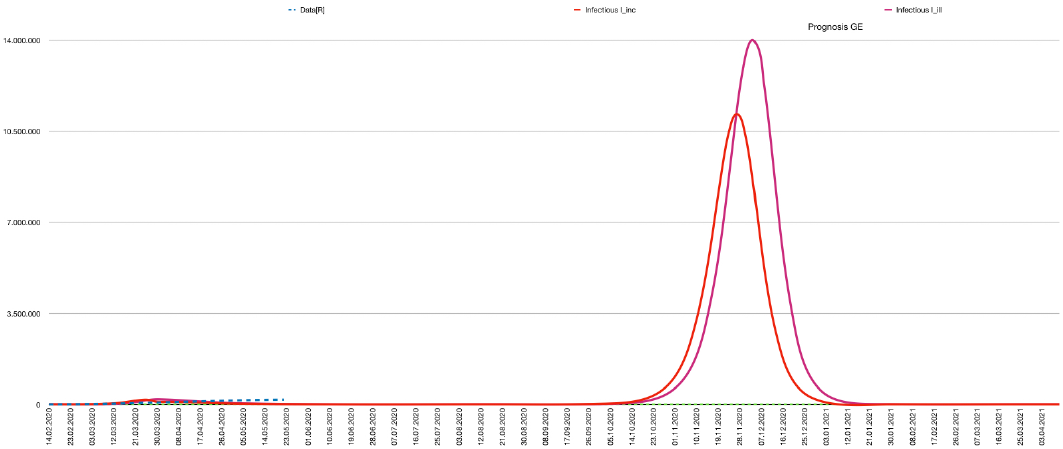
Emerging new massive wave due to incomplete extinction or introduction from abroad.

## 6. Discussion

To facilitate the discussion, we provide a rough evaluation scheme of the indicators and risks in the following table.

**Tab. 1.** Evaluation scheme.

| Indicator | RED | GREEN | Category | “green” Rationale |
| --- | --- | --- | --- | --- |
| Lifting | < 30 % | ≥ 30 % | Economy, Society | Significant increase of economical activities possible |
| PHC Load | >14.000 | ≤ 14.000 | Society | No „triage“ or rejections in hospitals due to ITM bottlenecks |
| Immunity | < 60 % | ≥ 60 % | Economy | No risk of severe „second wave“ |
| Deaths | > 40.000 | ≤ 40.000 | Society | No more death than in an Influenza epidemic |
| Date of Peak | 2021 | 2020 | Economy | Total lifting and thus full economic activity after peak date possible |
| Risk | Medium, High | Low |  | Probability of unintended second wave |

Using the evaluation scheme in Tab.1 the next table summarizes the findings of Chapter 6 for each scenario.

**Tab. 2.** Classification of Scenarios.

| Scenario | A: Leave-As-Is | B: Partial “green” Lifting | B: Partial “yellow” Lifting | C: Gradual “yellow” Lifting | D: Total Lifting | E: Shutdown |
| --- | --- | --- | --- | --- | --- | --- |
| Lifting ($R_{init}$) | 0% (0.75) | 18 % (0.99) | 68% (1.64) | 7 x 4.81% (1.19) | 100% (2.06) | -47% (0.0) |
| PHC Load | 630 | 630 | 14.000 | 14.000 | 30.000 | 630 |
| Immunity | 21 % | 25 % | 75 % | 67 % | 86 % | 21 % |
| Deaths | 10.400 | 40.000 | 380.000 | 330.000 | 480.000 | <10.000 |
| Peak | - | March 2021 | October 2020 | February 2021 | August 2020 | - |
| Risk | Low | Medium | Low | Low | Low | High |

Dependent on priority the scenarios can be roughly divided into a *social* (Scenario A, B-“green” and E) and an *economic* (Scenario B-“yellow”, C and D) group.

### Social Group

*Scenario A* maintains severe restrictions for a long period of time, till a vaccine is available. The death quote can be kept relatively low, but social upheavals would be possible.

*Scenario B-“green”* allows a small lifting of interventions in 2020 to calm down social conflicts and has an “acceptable” low death number comparable to other epidemics. The interventions have to be established till a vaccine is available. The price for the remaining long lasting interventions might be severe economical and social distortions. Furthermore, to mitigate the risk of missing the “green” corridor - besides trial-and error methods - the research on effects on lifting measures (simulations, etc.), have to be enforced.

*Scenario E* might be difficult to implement in an open society. As herd immunity will be not achieved at all, automated contact tracing might be mandatory to avoid a renewed infection wave.

### Economic Group

*Scenario D* will allow an early restart of economy after achieving herd immunity in September 2020, but with a very high loss of lives and situations, visible to society, much more worse than the “pictures in Italy and Spain”.

*Scenario B-yellow* has similar high losses of lives than Scenario D, but might avoid “cruel pictures” as Public Health Care would not be overloaded. Economy can fully recover in October 2020 when herd immunity is nearly reached.

Though allowing 35 % less overall lifting than Scenario B-yellow, *Scenario C* has similar characteristics for all indicators: high death quote, sufficient Public Health Care equipment, close to herd immunity. Full recovery of economy with some interventions in place (no *full* herd immunity) will be after February 2021. If no vaccine will be available in 2021, a second controllable gradual wave in 2021 might be necessary to gain full herd immunity. The risk of missing the 4,8% corridor per gradual lift, can be mitigated by controlling and adapting the effective R-value in succeeding intervention steps without overshooting of liftings (contrary to Scenario B-yellow).

## 7. Conclusion

Based on the notion of interventions-lifting, we defined six selected scenarios and four indicators spanning a decision space for non-medical interventions in the CoV-2 pandemic.

Using a calibrated simulation of an adopted SEIR-model we evaluated the scenarios against the indicators and classified them in a social and economic group.

Unfortunately, some of the parameters like the dark-figure, initial infections or initial population immunity are currently unknown, but have an significant effect on the predictions. Nevertheless, we tried to adapt the model to available data by selecting values for those parameters providing a simulation with high *R*^2^ (to current data), and good convergence of the measured and estimated *R*-value.

We found an anomaly of the ITM-function which allows an significant extension of intervention-liftings not overloading Public Health Care.

Generally, there is a very sensitive inverse relationship between public immunity and death numbers and due to the anomaly of the ITM/function a weaker one between public immunity and Public Health Care capacities.

In both groups the “Partial Lifting” Scenario B seems to have advantages over the other options. Nevertheless, due to the sensitivity of the number of deaths wrt. intervention/liftings at the “green/yellow” corridor-border there is a risk for the B-“green” lifting-strategy to end up in a region with high death numbers.

To meet and control this risk epidemiological and politically, may turn out to be very challenging.

## A. Model Equations

We use the definitions from Chapter 2. Actual data for each group **G** at day *n* is denoted by Data[**G**](*n*). The dynamic is modeled along Fig. 1 by the following system of entangled recursive (difference) equations.

I. Number of susceptible individuals **S**(*n*) where **I**(*n*) is defined in 5, is the difference between the population an the individuals who at time *n* have had already contact with the virus.

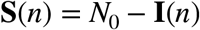
II. Daily increment of newly infected individuals Δ**I**(*n*) at time *n* is the part of susceptible individuals S(n) who are infected by infectious individuals **I***_inc_*(*n*) + **I***_ill_*(*n*) at time *n* via the daily effective replication rate *r_eff_* (*n*).

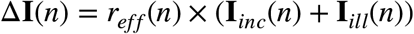
III. The number **E**(*n* + 1) of exposed, not infectious individuals at time *n* + 1 is the number **E**(*n*) of exposed individuals at time *n* incremented by the newly infected individuals Δ+**E**(*n*) at time *n*, and decremented by the infected individuals Δ^-^**E**(*n*) becoming infectious (and becoming not isolated) at time n after the latency time *λ*.

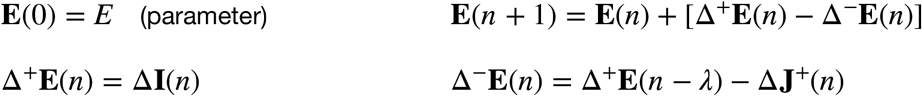
IV. Let *θ*(*n*): 0..1 define the ratio of isolated backtracked individuals to all backtracked individuals.The number of isolated individuals at time *n* + 1 (by automated contact backtracking) infected by individuals Δ^+^**R**(*n*) is

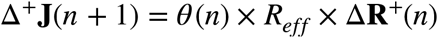 The part of isolated individuals getting ill and therefore registered at time *n* is

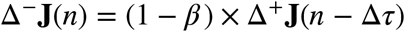
V. The number of infectious individuals within incubation time **I***_inc_*(*n* + 1) is the number of infectious individuals *I_inc_*(*n*) increased by the number of individuals Δ^+^**I***_inc_*(*n*) becoming infectious at time *n* and decreased by the number 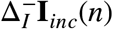 of individuals at time *n* becoming symptomatic after the incubation time and the number of isolated individuals 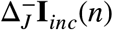.

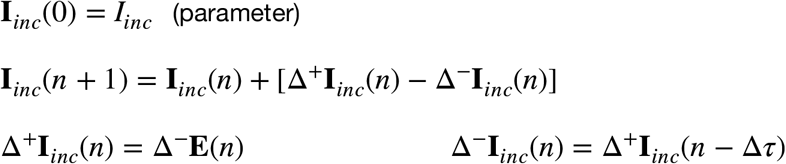
VI. The number of infectious (non registered) individuals with symptoms **I***_ill_*(*n* + 1) at time *n* is the number of infectious symptomatic unregistered individuals **I***_ill_*(*n*) incremented by the number Δ^+^**I***_ill_*(*n*) of individuals becoming weakly symptomatic without registration minus the number Δ^-^**I***_ill_*(*n*) of sick unregistered individuals becoming immune after the infectiousity time *σ_ill_*

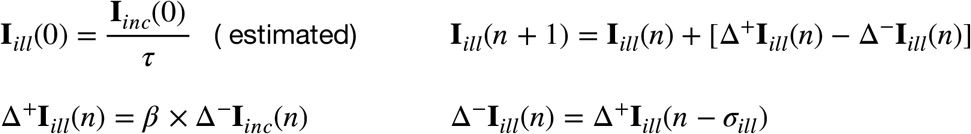
VII. Number of recovered (unregistered) individuals **N**(*n*) is increased by the number of recovered unregistered ill people.

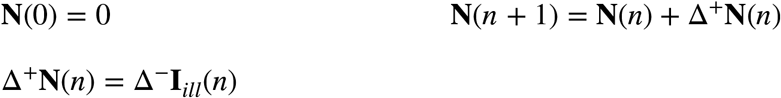
VIII. The accumulated number of noninfectious registered individuals **R**(*n* + 1) at time *n* + 1 is the number **R**(*n*) of registered symptomatic individuals at time *n* increased by the number Δ^+^**R**(*n*) of symptomatic or isolated individuals getting registered.

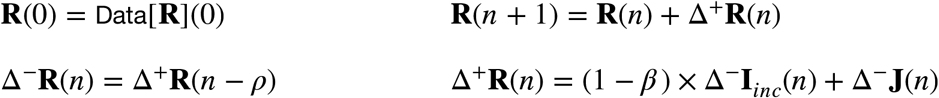
IX. The number of registered individuals **R***_itu_*(*n* + 1) requiring ITM at time *n* + 1 is the number of individuals **R***_itu_*(*n*) requiring ITM at time *n* increased by registered individuals Δ^+^*R_itu_*(*n*) needing ITM *ρ* days after getting symptomatic and decreased by the number Δ^-^**R***_itu_*(*n*) of individuals recovered from ITM or died after *ω* days.

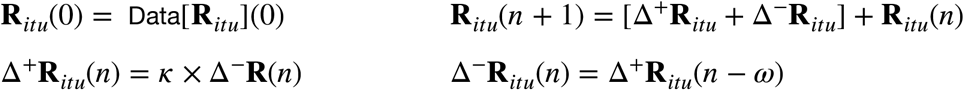
X. The aggregated number **D***_itu_*(*n* + 1) of deaths at time *n* + 1 caused by an ITM bottleneck is the number of deaths **D***_itu_*(*n*) at time *n* increased by deaths due to the missing ITM equipment **R***_itu_*(*n*) − *b* at day *n* where *b* is the number of maximal available ITM/RU units.

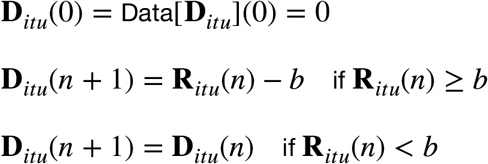
XI. The number of deaths **R***_dead_*(*n* + 1) at day *n* + 1 in spite of ITM is the number of deaths **R***_dead_*(*n)* at day *n* increased by the number Δ^+^**R***_dead_*(*n*) of deaths after ITM time *ω*.

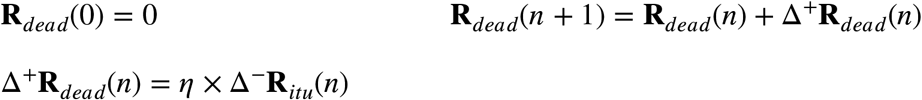
XII. The number of recovered registered individuals **R***_rec_*(*n* + 1) at day *n* + 1 is the number of recovered registered individuals **R***_rec_*(*n*) at time *n* incremented by the number Δ^+^**R***_rec_*(*n*) of recovered registered individuals Δ^+^**R***_rec_*(*n*) with or without ITM.

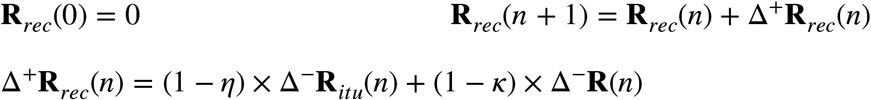
XIII. The aggregated number of infected, immune or isolated (non susceptible) individuals **I**(*n)* is

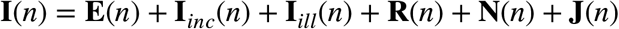

The following matrix gives an overview of the data flow between groups.

**Tab. 3.** Data flow between groups.

| to-><br>from | s | E | $\mathbf{I}_{inc}$ | $\mathbf{I}_{ill}$ | $\mathbf{R}$ | $\mathbf{R}_{itu}$ | $\mathbf{R}_{rec}$ | $\mathbf{R}_{dead}$ | N | J |
| --- | --- | --- | --- | --- | --- | --- | --- | --- | --- | --- |
| **S** | | $\Delta \mathbf{I}$ | | | | | | | | |
| **E** | | | $\Delta^- \mathbf{E}$ | | | | | | | $\Delta^+ \mathbf{J}$ |
| $\mathbf{I}_{inc}$ | | | | $\beta \times \Delta^- \mathbf{I}_{inc}$ | $(1 - \beta) \times \Delta^- \mathbf{I}_{inc}$ | | | | | |
| $\mathbf{I}_{ill}$ | | | | | | | | | $\Delta^- \mathbf{I}_{ill}$ | |
| **R** | | | | | | $\kappa \times \Delta^- \mathbf{R}$ | | | | |
| $\mathbf{R}_{itu}$ | | | | | | | $(1 - \eta) \times \Delta^- \mathbf{R}_{itu}$<br>$+ (1 - \kappa) \times \Delta^- \mathbf{R}$ | $\eta \times \Delta^- \mathbf{R}_{itu}$ | | |
| **J** | | | | | $\Delta^- \mathbf{J}$ | | | | | |

## B. Implementation

The recursive difference equation system has been implemented in IOS Numbers. Each line corresponds to one simulated day. The SEIR groups and their Δ-values are represented by corresponding columns, i.e. if *B* is the column representing **I**, then **I**(*n*) is implemented by the cell *B*(*n+”some Offset”)*.The implementation of the equations and parameters is straightforward.

## Data Availability

All Data referred in the document are available

## References

1 Hiroshi Nishiura & Gerardo Cowell. 2009. The Effective Reproduction Number as a Prelude to Statistical Estimation of Time-Dependent Epidemic Trends. Mathematical and Statistical Estimation Approaches in Epidemiology 103–119. doi: 10.1007/978-90-481-2313-15.

2 Mathias an der Heiden & Udo Buchholz. 2020. Modellierung von Beispielszenarien der SARS-CoV-2-Epidemie 2020 in Deutschland. https://www.rki.de/DE/Content/InfAZ/NZNeuartiges_Coronavirus/Modellierung_Deutschland.pdf?___blob=publicationFile

3 2020. SARS-CoV-2 Steckbrief zur Coronavirus-Krankheit-2019 (COVID-19). Robert Koch Institut Website: Robert Koch Institut. https://www.rki.de/DE/Content/InfAZ/N/Neuartiges_Coronavirus/Steckbrief.html

4 Matthias an der Heiden & Osama Hamouda. 2020. Schätzung der aktuellen Entwicklung der SARS-CoV-2 Epidemie in Deutschland - Nowcasting. Robert Koch Institut.

5 CSSE John Hopkins University. 2020. COVID-19 Dashboard. https://www.arcgis.com/apps/opsdashboard/index.html#/bda7594740fd40299423467b48e9ecf6

6 Deutsche Krankenhausgesellschaft. 2020. Coronavirus: Fakten und Infos. Deutsche Krankenhausgesellschaft.

